# Sleep duration irregularity and risk for incident cardiovascular disease in the UK Biobank

**DOI:** 10.1101/2024.07.26.24311090

**Authors:** Tianyi Huang, Sina Kianersi, Heming Wang, Kaitlin S. Potts, Raymond Noordam, Tamar Sofer, Martin K. Rutter, Susan Redline

## Abstract

**Background:** Emerging evidence supports a link between circadian disruption as measured by higher night-to-night variation in sleep duration and increased risk of cardiovascular disease (CVD). It remains unclear whether this association varies by CVD types or may be modified by average sleep duration and genetic risk for CVD.

**Methods:** Our prospective analysis included 86,219 UK Biobank participants who were free from CVD when completing 7 days of accelerometer measurement in 2013-2016. Sleep irregularity was evaluated by the standard deviation (SD) of accelerometer-measured sleep duration over 7 days. Incident major CVD events, defined as fatal or nonfatal myocardial infarction (MI) and stroke, were identified through linkage to Hospital Episode Statistics data until May 31, 2022. Multivariable-adjusted Cox proportional hazard models were used to estimate hazard ratios (HRs) and 95% CIs for associations of sleep duration SD with risk for major CVD events overall and for MI and stroke separately.

**Results:** We documented 2,310 incident cases of major CVD events (MI: 1,183, stroke: 1,175) over 636,258 person-years of follow-up. After adjusting for sociodemographic factors and family history of CVD, the HR (95% CI) associated with a 1-hour increase in sleep duration SD was 1.19 (1.10, 1.27) for CVD (p-trend<0.0001), 1.23 (1.11, 1.35) for MI (p-trend<0.0001), and 1.17 (1.05, 1.29) for stroke (p-trend=0.003). Additional adjustment for lifestyle factors, co-morbidities and sleep-related factors modestly attenuated these associations. Higher sleep irregularity was associated with higher CVD risk irrespective of genetic risk (p-interaction=0.43), but this association was stronger among individuals with longer average sleep duration >8 hours (p-interaction=0.006)

**Conclusions:** Higher night-to-night variation in accelerometer-measured sleep duration was associated with consistently higher risks for major CVD events. The association did not seem to be modified by genetic risk for CVD and was more pronounced in long sleepers.

## Introduction

Sleep health is increasingly recognized as a crucial component of a healthy lifestyle especially for promoting cardiovascular well-being. Numerous prospective epidemiologic studies have linked self-reported short and long sleep duration with higher cardiovascular disease (CVD) incidence and mortality.^1^ Recent results based on objective sleep assessment or Mendelian randomization also support increased CVD risk and mortality associated with short sleep, although the evidence for long sleep duration is weaker and less consistent.^2–4^ Built upon the mounting evidence, the American Heart Association in 2022 updated the construct of cardiovascular health from Life’s Simple 7 to Life’s Essential 8, highlighting the role of sleep health in CVD prevention and focusing on advocating 7-8 hours of habitual sleep.^5^

Notably, the mean level of sleep duration, either estimated by self-reports or averaged across multiple nights of objective monitoring, only captures one aspect of sleep health.^6,7^ Maintaining consistent sleep-wake cycles is essential for achieving restful, restorative sleep and synchronizing internal circadian rhythms with the social and natural environment. Growing evidence suggests that night-to-night variability in sleep duration, independent of mean sleep duration, is a novel risk factor for cardiometabolic disease and mortality.^8–12^ In the Multi-Ethnic Study of Atherosclerosis (MESA), we reported that higher sleep duration irregularity measured by 7-day standard deviation (SD) in actigraphy-based sleep duration was significantly associated with greater subclinical atherosclerotic burden and higher CVD risk after accounting for traditional CVD risk factors and average sleep duration.^8,9^ However, the modest sample size in MESA constrained our ability to fully explore potential heterogeneity in the association between sleep duration irregularity and CVD risk by major CVD types. It also remains unclear whether sleep duration irregularity, a behavior amenable to modification, may interact with genetic susceptibility to CVD or average sleep duration to influence CVD risk.

Leveraging the large-scale accelerometer-measured sleep data from the UK Biobank, we evaluated the prospective associations of 7-day variability in sleep duration with incidence of major adverse cardiovascular events overall and separately for myocardial infarction (MI) and stroke. We hypothesized that higher variability in sleep duration would exacerbate CVD risk similarly for MI and stroke, with stronger associations in individuals with a greater genetic predisposition to CVD or shorter average sleep duration.

## Methods

### Study population

The UK Biobank is a large prospective cohort study initiated in 2006-2010, when over 500,000 participants aged 40-69 years completed baseline physical, sociodemographic, and medical assessments in one of 22 centers in the UK.^13^ Participants were prospectively followed for occurrence of a wide range of health outcomes, with collection of additional exposure and phenotypic information through further assessment. In 2013-2015, a subsample of 103,712 participants completed an accelerometer study over a 7-day period.^14^ Our analysis included 92,884 participants who provided ≥5 days of accelerometer sleep data. Of these, we further excluded participants with indicators for poor data quality (e.g., insufficient wear time; n=3,355), extreme sleep values (e.g., average sleep duration <3 hours or >12 hours or sleep duration SD >4 hours; n=114), pre-existing CVD prior to the end of accelerometer assessment (n=3,193), or loss to follow-up (n=3), leaving 86,219 for analysis. The UK Biobank received approval by the NHS North-West Multi-Centre Research Ethics Committee. All participants provided written informed consent. The present study was completed under the UK Biobank project 85501.

### Sleep assessment

Between 2013-2015, participants who provided a valid email address were invited to wear a wrist accelerometer (Axivity AX3) for 7 days.^14^ A total of 103,712 participants returned the data, which were further processed by the R package GGIR consistent with prior research.^15,16^ Sleep periods and episodes were identified based on 5-second epoch time-series accelerometer signals, and determined as the longest block of combined inactivity bouts lasting ≥30 min. This algorithm has been validated against polysomnography to accurately detect sleep periods in the absence of sleep diary records.^17^ For each participant, we calculated the mean and SD of sleep duration across all available days, and the sleep duration SD was used as the measure for sleep duration irregularity. Sleep efficiency was defined as proportion of sleep duration within the sleep period window (i.e., the time between the start and end of the longest inactivity block).

At UK Biobank enrollment (2.8 to 9.7 years prior to the accelerometer study), participants also self-reported several habitual sleep symptoms or patterns, with each assessed using a single, representative question carefully chosen from commonly used instruments. These sleep metrics included sleep duration, insomnia symptoms, chronotype, snoring and daytime sleepiness.^18^ Sleep apnea was identified through a combination of self-reported diagnosis and medical records.^19^

### Ascertainment of CVD events

Incidence of major CVD events was defined as the first occurrence of MI or stroke that resulted in hospitalization or death. Hospital admissions were identified through linkage to the Hospital Episode Statistics, and date of death was identified based on death certificates from the National Death Register. Classification of fatal and nonfatal CVD events was recorded using the International Classification of Diseases, Tenth Revision (ICD-10). We considered ICD-10 codes I21-23 for MI and I60-64 for stroke.

### Statistical analysis

For each participant, we calculated person-time of follow-up from the end date of the accelerometer study until the earliest date of major CVD events, death, or end of follow-up (May 31, 2022). Cox proportional hazards regression was used to estimate the association between sleep duration SD and risk of developing CVD events over follow-up. We did not detect significant deviation from the proportional hazards assumption by testing the time-by-exposure interaction (p for interaction=0.36). Sleep duration SD was modeled as a continuous variable (per 1-hour) as well as in categories (≤30, 31-45, 46-60, 61-90, >90 min). The primary model adjusted for age, sex, self-reported race and ethnicity, Townsend deprivation index, work schedules, and family history of CVD. Lifestyle, co-morbidities and sleep disturbances may be consequences of irregular sleep duration, therefore acting in the pathway linking sleep duration SD and CVD risk. To avoid potential overadjustment,^20^ we considered these covariates in separate multivariable models. In the second model, we further controlled for BMI, smoking status, alcohol consumption, diet quality, physical activity, and history of hypertension, dyslipidemia, diabetes and depression. We additionally added to the third model objectively-measured average sleep duration and sleep efficiency, self-reported insomnia symptoms, chronotype and sleepiness, and clinically diagnosed sleep apnea. All analyses were performed for major CVD events overall and for MI and stroke separately.

To test the robustness of the results, we performed several sensitivity analyses. First, given that the COVID-19 pandemic may have disrupted healthcare, impacting cardiovascular testing and diagnosis and resulting in outcome misclassification and potentially biased associations,^21^ we repeated the analysis with the end of follow-up as March 23, 2020, the start of the UK national lockdown. Second, we excluded CVD events occurring within the first year after the accelerometer study to assess the impact of preclinical CVD and symptoms on sleep duration irregularity (i.e., reverse causation). Third, considering that variation in sleep schedules between weekdays versus weekend (e.g., weekend catch-up sleep) is an important contributor to day-to-day variability in sleep duration across the week ^22^, we assessed whether the observed associations for sleep duration irregularity persisted beyond weekday-weekend variation by focusing on sleep duration SD across weekdays only. Fourth, we examined the associations separately for ischemic and hemorrhagic strokes, as these stroke subtypes may have distinct etiologic origins.^23^

We conducted subgroup analysis by age, sex, race, average sleep duration, polygenic risk score (PRS) for CVD, and family history of CVD based on the primary multivariable model. Subgroup heterogeneity was evaluated by the likelihood ratio test of the multiplicative interaction with the continuous sleep duration SD. Of *a priori* interest, we examined the joint associations between sleep duration SD (≤30, 31-45, 46-60, 61-90, >90 min) and PRS (low, medium, high) or average sleep duration (<7, 7-8, >8 hrs) with CVD outcomes. Participants with sleep duration SD <30 min and low PRS or 7-8 hrs of average sleep duration were used as the reference group in the joint analyses. All statistical analyses were performed using SAS version 9.4 (SAS Institute).

## Results

Of 86,219 participants, the mean (SD) for 7-day SD in sleep duration was 56 (33) minutes, with 25,434 (29.5%) having sleep duration SD>60 minutes. Participants with higher sleep duration irregularity, assessed by sleep duration SD, tended to be younger, female, non-white, socioeconomically disadvantaged and employed, especially in occupations requiring shift work, and were more likely to have higher BMI and currently smoke (**Table 1**). Family history of CVD was slightly less common in participants with higher sleep duration SD, but no clear difference was observed when genetic risk for CVD was assessed by PRS. Although the prevalence of metabolic disorders (dyslipidemia, hypertension, diabetes) did not differ appreciably across categories of sleep duration SD, the prevalence of depression was higher among participants with greater sleep duration SD. With increasing sleep duration SD, there was a notable trend towards shorter objective sleep duration, particularly among those with sleep duration SD≥90 minutes. Additionally, irregular sleepers exhibited lower sleep efficiency and were more likely to self-report as evening chronotypes.

**Table 1.**
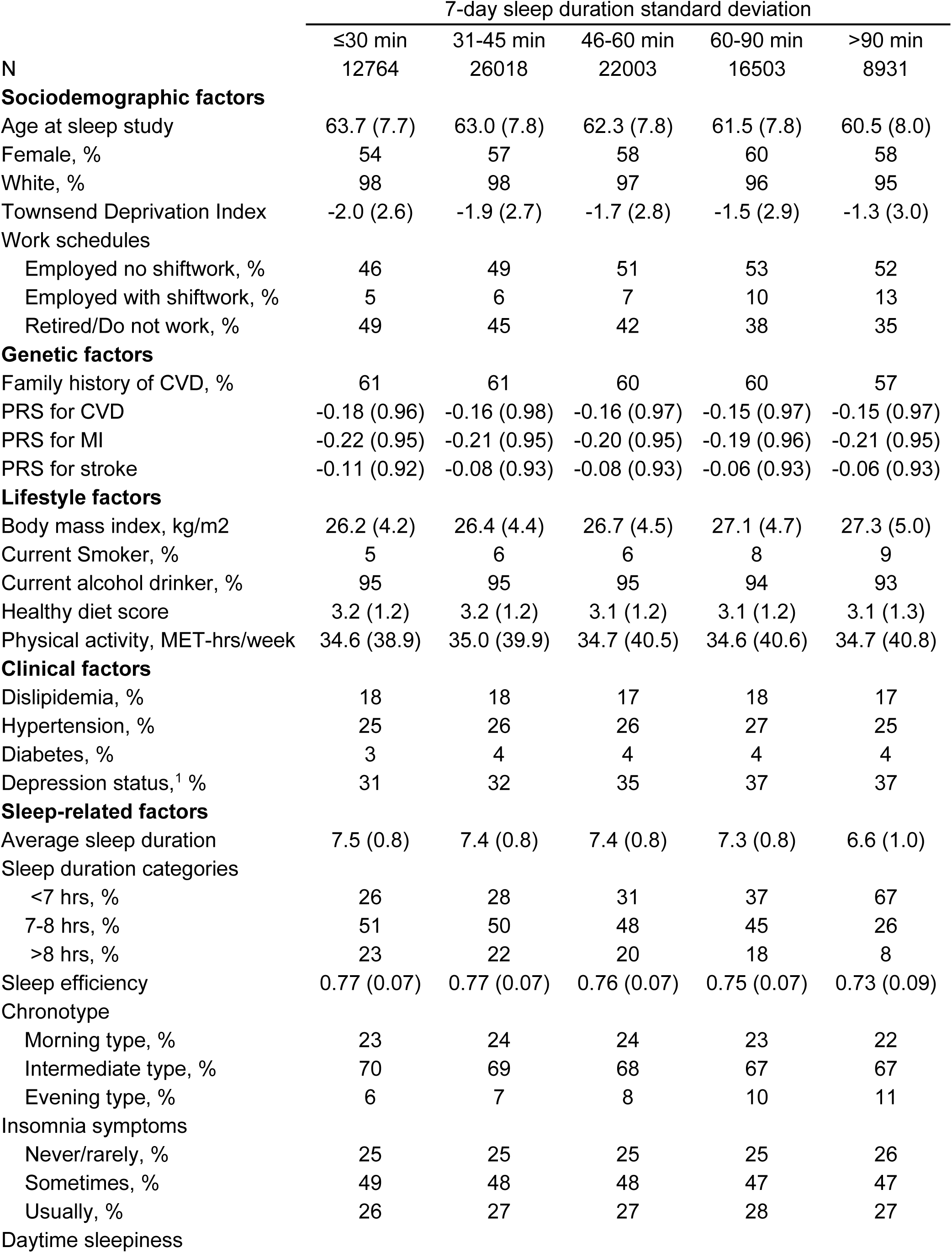
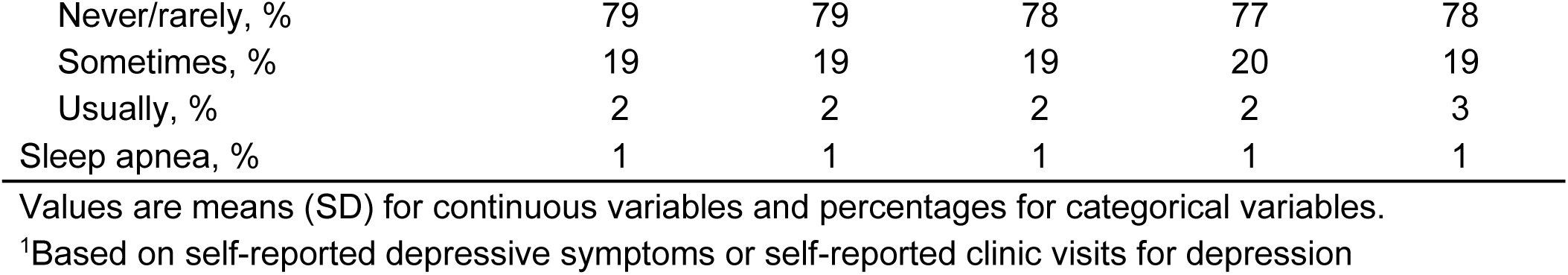
Characteristics of the study population by category of sleep duration irregularity.

### Associations of sleep duration irregularity and cardiovascular disease risk

Over 636,258 person-years of follow-up (median: 7.5 years), 2,310 participants developed a major CVD event (1,183 MI and 1,175 stroke). After adjusting for sociodemographic factors and family history of CVD, the HR (95% CI) for major CVD events was 1.10 (0.97, 1.26) for those with a sleep duration SD of 31-45 minutes, 1.19 (1.04, 1.37) for 46-60 minutes, 1.33 (1.15, 1.53) for 61-90 minutes, and 1.40 (1.18, 1.65) for >90 minutes, compared with participants with a sleep duration SD ≤30 minutes (p for trend<0.0001; **Table 2**). Similar positive associations were observed for MI and stroke separately. The HR (95% CI) associated with a 1-hour increase in sleep duration SD was 1.19 (1.10, 1.27) for CVD, 1.23 (1.11, 1.35) for MI, and 1.17 (1.05, 1.29) for stroke. Further adjustment for lifestyle factors, co-morbidities and sleep-related factors only modestly attenuated the associations. For example, the HR (95% CI) for major CVD events per 1-hour increment in sleep duration SD was 1.15 (1.07, 1.24) after further adjusting for lifestyle factors and co-morbidities (p for trend=0.0001) and 1.13 (1.05, 1.22) after additionally controlling for sleep-related factors (p for trend=0.001).

**Table 2.** Associations of accelerometer-measured sleep duration irregularity with incident cardiovascular events in the UK Biobank.

|  | 7-day sleep duration standard deviation |  |  |  |  | Per 1 hour | P-trend |
| --- | --- | --- | --- | --- | --- | --- | --- |
|  | ≤30 min | 31-45 min | 46-60 min | 60-90 min | >90 min |  |  |
| Major cardiovascular events |  |  |  |  |  |  |  |
| Cases | 327 | 691 | 589 | 455 | 248 | 2310 |  |
| Person-years | 94088 | 192004 | 162509 | 121711 | 65945 | 636258 |  |
| Model 1 | Ref | 1.10 (0.97, 1.26) | 1.19 (1.04, 1.37) | 1.33 (1.15, 1.53) | 1.40 (1.18, 1.65) | 1.19 (1.10, 1.27) | <.0001 |
| Model 2 | Ref | 1.09 (0.95, 1.24) | 1.15 (1.01, 1.32) | 1.25 (1.08, 1.44) | 1.31 (1.11, 1.55) | 1.15 (1.07, 1.24) | 0.0001 |
| Model 3 | Ref | 1.09 (0.95, 1.24) | 1.15 (1.00, 1.31) | 1.24 (1.07, 1.43) | 1.27 (1.07, 1.50) | 1.13 (1.05, 1.22) | 0.001 |
| Myocardial infarction |  |  |  |  |  |  |  |
| Cases | 176 | 331 | 310 | 228 | 138 | 1183 |  |
| Person-years | 94454 | 192993 | 163280 | 122333 | 66255 | 639316 |  |
| Model 1 | Ref | 0.98 (0.82, 1.18) | 1.16 (0.96, 1.40) | 1.21 (0.99, 1.48) | 1.39 (1.11, 1.74) | 1.23 (1.11, 1.35) | <.0001 |
| Model 2 | Ref | 0.96 (0.80, 1.15) | 1.11 (0.92, 1.33) | 1.12 (0.92, 1.36) | 1.28 (1.02, 1.60) | 1.18 (1.07, 1.31) | 0.001 |
| Model 3 | Ref | 0.96 (0.80, 1.15) | 1.09 (0.91, 1.31) | 1.10 (0.90, 1.34) | 1.20 (0.96, 1.51) | 1.15 (1.04, 1.27) | 0.008 |
| Stroke |  |  |  |  |  |  |  |
| Cases | 159 | 369 | 290 | 237 | 120 | 1175 |  |
| Person-years | 94519 | 192940 | 163443 | 122385 | 66379 | 639665 |  |
| Model 1 | Ref | 1.21 (1.01, 1.46) | 1.21 (1.00, 1.47) | 1.44 (1.18, 1.77) | 1.43 (1.13, 1.82) | 1.17 (1.05, 1.29) | 0.003 |
| Model 2 | Ref | 1.20 (1.00, 1.45) | 1.19 (0.98, 1.44) | 1.38 (1.13, 1.70) | 1.38 (1.08, 1.75) | 1.15 (1.03, 1.27) | 0.01 |
| Model 3 | Ref | 1.20 (1.00, 1.45) | 1.19 (0.98, 1.44) | 1.38 (1.13, 1.70) | 1.36 (1.07, 1.74) | 1.14 (1.03, 1.27) | 0.01 |

Model 1: adjusted for age, sex, race, Townsend deprivation index, work schedules, and family history of CVD
Model 2: Model 1 + adjusted for BMI, smoking status, alcohol consumption, diet quality, physical activity, and history of hypertension, dyslipidemia, diabetes and depression
Model 3: Model 2 + adjusted for accelerometer-measured average sleep duration and sleep efficiency, self-reported insomnia symptoms, chronotype and daytime sleepiness, and clinically diagnosed sleep apnea based on ICD-10

### Sensitivity analyses

We observed somewhat stronger associations when the analysis was censored at the start of the UK COVID-19 lockdown (**Supplemental Table 1**). For example, the HR (95% CI) for CVD after adjustment for sociodemographic factors and family history of CVD was 1.15 (0.98, 1.36) for sleep duration SD of 31-45 minutes, 1.28 (1.08, 1.51) for 46-60 minutes, 1.42 (1.19, 1.69) for 61-90 minutes, and 1.51 (1.23, 1.85) for >90 minutes, compared with sleep duration SD ≤30 minutes (p for trend<0.0001). Slightly weaker but significant positive associations were observed in sensitivity analyses excluding CVD events diagnosed within the first year after the sleep assessment (**Supplemental Table 2**) or only evaluating sleep duration variability across weekdays (**Supplemental Table 3**). When examining stroke risk by type, the higher stroke risk associated with irregular sleep duration was primarily attributed to ischemic stroke rather than hemorrhagic stroke (**Supplemental Table 4**).

### Subgroup associations

The higher CVD risk associated with irregular sleep duration was more pronounced among participants ≥60 years versus <60 years (p-interaction=0.04) and among participants with longer average sleep duration >8 hours (p-interaction=0.006; **Table 3**). We observed a stronger positive association between sleep duration irregularity and CVD risk among participants with a family history of CVD compared to those without the history (p-interaction=0.03). However, such potential gene-sleep interactions were not observed when genetic predisposition was assessed by PRS (p-interaction=0.43), and irregular sleep duration was associated with higher CVD risk regardless of the PRS for CVD. There was no significant difference in the association by sex (p-interaction=0.09) or race (p-interaction=0.42), although the race-stratified analysis was underpowered. These subgroup differences remained largely the same when MI and stroke were examined individually, except that the association for stroke did not differ by family history of CVD and was significantly stronger in men than in women **(Supplemental Table 5).**

**Table 3.** Subgroup analysis of the association between accelerometer-measured sleep duration irregularity and cardiovascular disease risk in the UK Biobank.

|  | Cases/N | HR (95% CI)<sup>1</sup> | P-int |
| --- | --- | --- | --- |
| Age |  |  | 0.04 |
| <60 yrs | 393/31903 | 1.02 (0.86, 1.20) |  |
| ≥60 yrs | 1917/54316 | 1.23 (1.13, 1.33) |  |
| Sex |  |  | 0.09 |
| Men | 1414/36651 | 1.22 (1.11, 1.33) |  |
| Women | 896/49568 | 1.13 (1.00, 1.27) |  |
| Race |  |  | 0.42 |
| White | 2242/83577 | 1.19 (1.11, 1.28) |  |
| Non-white | 68/2642 | 1.01 (0.68, 1.51) |  |
| Average sleep duration |  |  | 0.006 |
| <7 hrs | 876/29488 | 1.07 (0.97, 1.18) |  |
| 7-8 hrs | 983/39890 | 1.22 (1.07, 1.40) |  |
| >8 hrs | 451/16841 | 1.52 (1.23, 1.89) |  |
| Family history of CVD |  |  | 0.03 |
| No | 759/34288 | 1.07 (0.94, 1.22) |  |
| Yes | 1551/51931 | 1.24 (1.14, 1.35) |  |
| PRS<sup>2</sup> |  |  | 0.43 |
| Low | 544/28033 | 1.17 (1.01, 1.36) |  |
| Medium | 721/28034 | 1.34 (1.19, 1.51) |  |
| High | 991/28034 | 1.09 (0.97, 1.22) |  |

<sup>1</sup>Adjusted for age, sex, race, Townsend deprivation index, work schedules, and family history of CVD
<sup>2</sup>Among 84101 participants with genotyping information with additional adjustment of the first three genetic principal components

### Joint associations

Irregular sleep duration was consistently associated with increases in CVD risk across tertiles of PRS for CVD (**Figure 1a**). Compared with individuals with low genetic risk and a sleep duration SD≤30 min, the highest CVD risks were observed in individuals with high genetic risk and a sleep duration SD 61-90 minutes (HR: 2.52; 95% CI: 1.94, 3.28) or >90 minutes (HR: 2.39; 95% CI: 1.78, 3.22). Further, higher risks for MI or stroke associated with irregular sleep duration were observed across various genetic risk categories for MI or stroke, except for individuals with low genetic risk for stroke, where no clear association was found (**Supplemental Figure 1**). Compared with individuals with a sleep duration SD ≤30 min and an optimal sleep duration of 7-8 hours, the elevated CVD risk associated with a large sleep duration SD >90 minutes was more pronounced in long sleepers >8 hrs (HR: 1.95; 95% CI: 1.29, 2.93) than in short sleepers <7 hrs (HR: 1.42; 95% CI: 1.15, 1.77; **Figure 1b**). The joint association patterns were similar for MI and stroke individually (**Supplemental Figure 2**).

**Figure 1.**
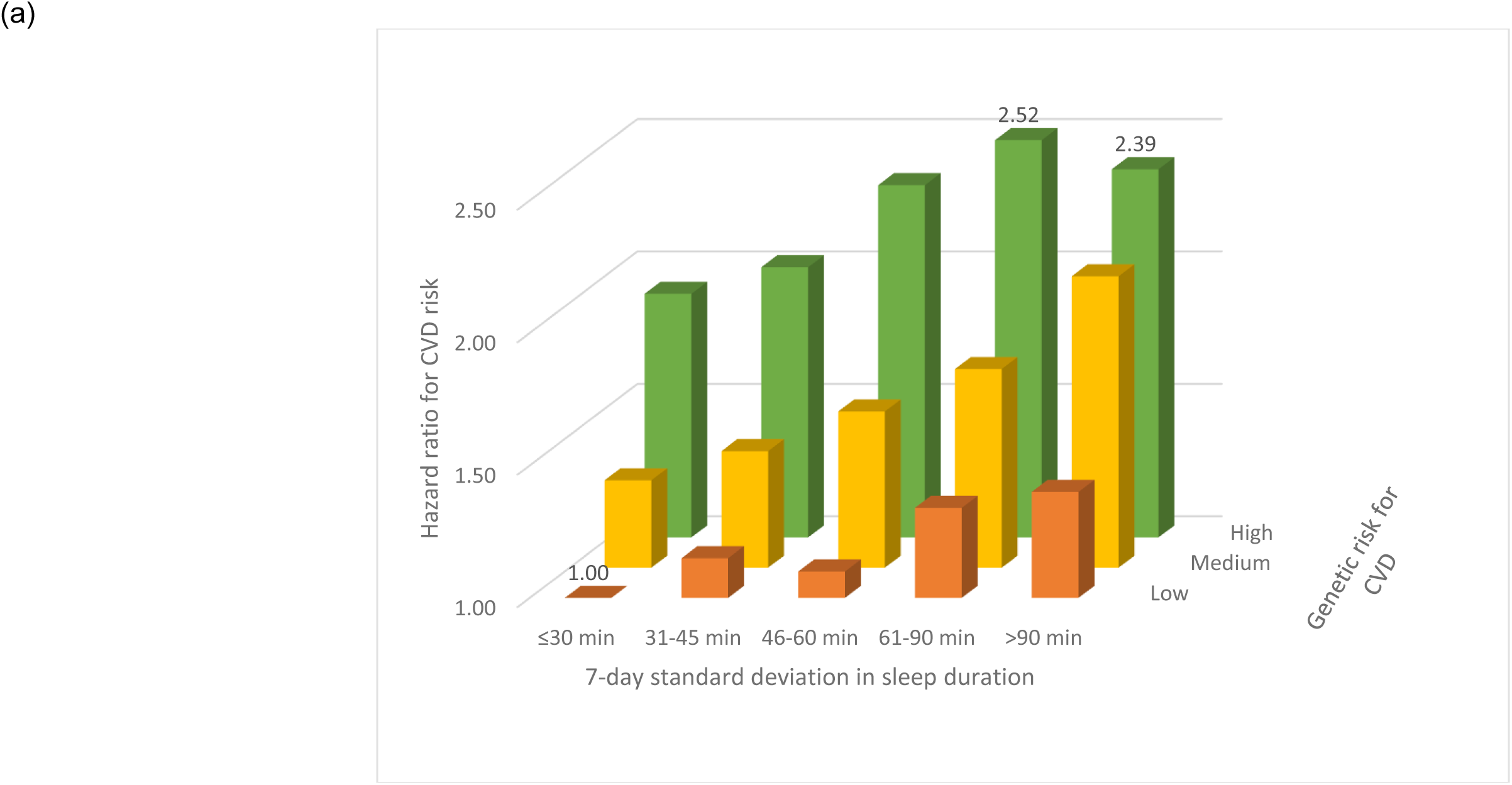
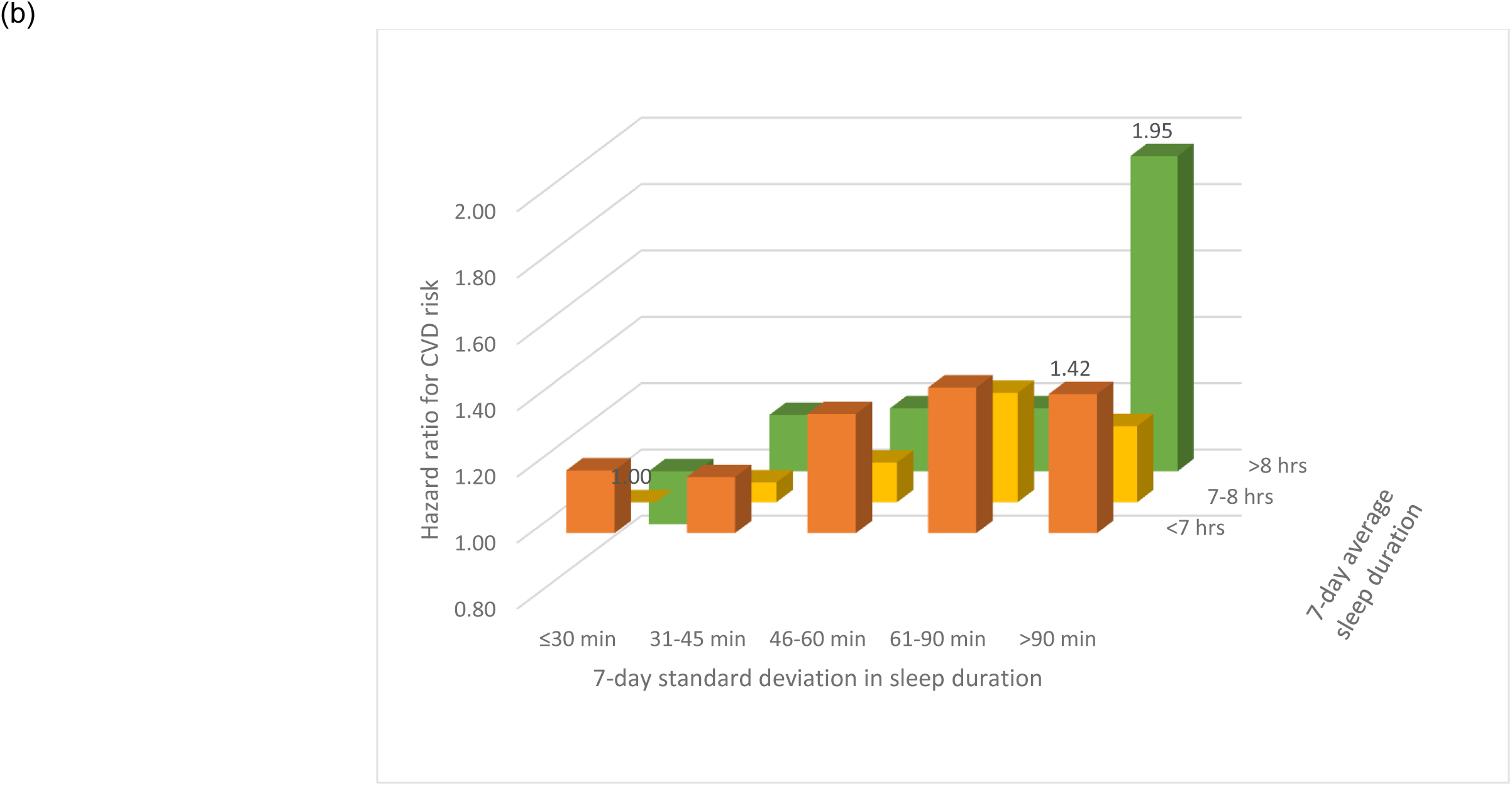
Risk of incident cardiovascular events according to joint categories of sleep duration irregularity and. (a) polygenic risk score for CVD and (b) average sleep duration. Estimates adjusted for age, sex, race, Townsend deprivation index, work schedules, and family history of CVD (plus the first three genetic principal components for joint associations with the polygenic risk score).

## Discussion

This prospective study in the UK Biobank shows higher CVD risks among individuals with irregular sleep duration after accounting for established CVD risk factors. The observed increase in CVD risk associated with irregular sleep duration was similar for MI and stroke. Although irregular sleep duration was consistently associated with higher CVD risk irrespective of the genetic risk profile or the average sleep duration, the association was more pronounced among individuals with sleep duration longer than 8 hours.

Consistent with prior studies linking sleep irregularity with higher risks for CVD^8^ and total mortality^10–12^ as well as with CVD risk factors including atherosclerotic development,^9^ metabolic syndrome^24^ and diabetes,^25^ our findings provide further evidence supporting the congruent associations of irregular sleep duration with risk of MI and stroke, particularly ischemic stroke. This suggests that irregular sleep duration may modulate common pathways leading to ischemic cardiovascular events. These pathways include dysregulation of blood pressure, glucose/lipid metabolism, inflammation, endothelial function, and thrombotic events induced by circadian disruption,^26–31^ as well as adverse effects of intermittent sleep curtailment on sympathetic and hormonal activities.^32–34^ Given that irregular sleep duration can occur on a daily, habitual basis, the resulting biologic changes may act as triggers for adverse cardiovascular events, significantly increasing the chance of developing CVD events over the long term. More direct evidence is needed to fully understand the mechanisms underlying the association between irregular sleep duration and CVD risk.

Of note, the association observed in the current study was more modest compared to our prior investigation in MESA.^8^ Specifically, the HR (95% CI) for CVD was 1.19 (1.10, 1.27) in the UK Biobank and 1.41 (1.12, 1.79) in MESA. While the smaller sample size in MESA may contribute to less accurate estimates, key differences between the cohorts could also explain this. For example, MESA included older individuals (mean age: 69 vs 62 years), and our subgroup analysis suggests that older individuals may be more vulnerable to the negative cardiovascular effects of irregular sleep duration. The prevalence of irregular sleep duration also differed, with 39.5% in MESA having a 7-day sleep duration SD >90 min compared to 10.4% in the UK Biobank. Further, the UK Biobank analysis included the COVID-19 period, which, according to sensitivity analysis, led to a weaker overall association. Greater misclassification of CVD outcomes due to delayed or missed diagnostic testing during the COVID-19 period,^21^ as well as inaccuracy of using data from 2013-2015 to reflect pandemic sleep duration irregularity when many were staying at home, may have both attenuated the observed association.

Consistent with our initial hypothesis, irregular sleep duration was more strongly associated with CVD risk among individuals with a family history of CVD, an indirect measure for inherited susceptibility. However, such a gene-sleep interaction was not present when PRS was used as the direct, quantitative measure for genetic susceptibility. A positive association between irregular sleep duration and CVD risk was consistently observed in individuals with low, medium, or high genetic risk, similar to a previous UK Biobank study evaluating gene-sleep interactions for CVD risk based on self-reported sleep traits.^18^ The inconsistent findings may be attributed to the fact that PRS and family history represent independent and complementary rather than interchangeable measures for inherited disease susceptibility.^35^ Family history encompasses additional lifestyle and environmental factors shared within families that may influence the associations beyond genetic susceptibility. It is also possible that irregular sleep duration may interact with specific genetic pathways to influence CVD risk. Significant gene-sleep interactions may be more likely to be observed with pathway-specific PRS rather than the overall CVD PRS,^19^ which requires future investigation.

Intriguingly, although shorter sleepers (<7 hours) tended to exhibit greater sleep duration irregularity, longer sleepers (>8 hours) seemed more susceptible to the adverse effects of sleep duration irregularity on CVD risk. This suggests that excessive catch-up sleep, which resulted in both longer average sleep duration and larger variability, may not compensate for sleep debt but conversely may lead to circadian dysregulation and increased CVD risk.^36^ Previous research on weekday-weekend differences in sleep duration only found beneficial relationships of shorter, but not longer, weekend catch-up sleep with inflammation and mortality.^37,38^ Our joint analysis suggests that irregular sleep duration coupled with long sleep duration conferred the highest CVD risk. Other studies reported that sleep irregularity was a stronger predictor of mortality than either sleep duration^11^ or sleep apnea.^12^ As a result, a recent consensus statement from the National Sleep Foundation highlighted the importance of sleep regularity for health and performance,^39^ calling for increased research and public initiatives to optimize sleep regularity at the population level.

The strengths of the study included the prospective design, large-scale accelerometer measurements, systematic identification of incident CVD outcomes through ICD-coded health records, and the availability of important covariates including genetic risk. However, the analytical sample from the UK Biobank included primarily white British participants with relatively high socioeconomic background, potentially limiting the generalizability of our findings to other populations. Nonetheless, the consistency with results from the racial and ethnic diverse MESA cohort^8^ suggests that the impact of irregular sleep duration on CVD risk may have broader implications. We relied on accelerometer measurement taken over 7 days to quantify irregular sleep duration, which may not reliably represent long-term habitual sleep patterns that are relevant for CVD pathogenesis, particularly during the pandemic. This limitation could weaken the observed associations and may partially explain why stronger associations were observed when the COVID-19 period was excluded from the analysis. Despite the possibility for residual confounding given that many covariates were assessed at the UK Biobank baseline 2.8 to 9.7 years prior to the accelerometer study, it is unlikely that misclassification of time-varying covariates (e.g., work schedules) could fully account for the associations observed. Finally, our study was observational by nature, and further evidence is needed to establish the causal relationship between irregular sleep duration and CVD risk.

In summary, irregular sleep duration as characterized by high day-to-day variation in habitual sleep duration over a 7-day period was associated with higher CVD risk overall as well as higher risk of MI and stroke individually in the UK Biobank. The association remained robust after adjustment for a wide variety of factors, did not differ appreciably by genetic risk for CVD, and was particularly strong in long sleepers and older individuals. These findings underscore the importance of maintaining consistent sleep duration for cardiovascular health and suggest that promoting regular sleep duration may be a novel behavioral strategy for CVD risk reduction that should be tested in intervention studies.

## Data Availability

All data and codes referred to in the manuscript are available upon request to the authors.

## Acknowledgements

This research was supported by NIH grant R01HL155395, UK Biobank project 85501 and the Intramural Research Program of the NIH, National Institute on Aging. SK was supported by the American Heart Association Postdoctoral Fellowship (24POST1188091).

---

**Supplemental Table 1.** Associations of accelerometer-measured sleep duration irregularity with incident cardiovascular events in the UK Biobank, censored at the start of the COVID-19 pandemic lockdown (March 23, 2020)

|  | 7-day sleep duration standard deviation |  |  |  |  | Per 1 hour | P-trend |
| --- | --- | --- | --- | --- | --- | --- | --- |
|  | ≤30 min | 31-45 min | 46-60 min | 60-90 min | >90 min |  |  |
| Major cardiovascular events |  |  |  |  |  |  |  |
| Cases | 205 | 449 | 393 | 304 | 173 | 1524 |  |
| Person-years | 69140 | 140444 | 119186 | 89709 | 50157 | 468635 |  |
| Model 1 | Ref | 1.15 (0.98, 1.36) | 1.28 (1.08, 1.51) | 1.42 (1.19, 1.69) | 1.51 (1.23, 1.85) | 1.24 (1.13, 1.35) | <.0001 |
| Model 2 | Ref | 1.13 (0.96, 1.34) | 1.23 (1.04, 1.46) | 1.32 (1.11, 1.58) | 1.41 (1.15, 1.72) | 1.20 (1.10, 1.31) | <.0001 |
| Model 3 | Ref | 1.13 (0.96, 1.33) | 1.22 (1.03, 1.45) | 1.31 (1.09, 1.57) | 1.35 (1.10, 1.67) | 1.17 (1.07, 1.28) | 0.0008 |
| Myocardial infarction |  |  |  |  |  |  |  |
| Cases | 109 | 218 | 213 | 160 | 97 | 797 |  |
| Person-years | 69323 | 140940 | 119576 | 90017 | 50329 | 470185 |  |
| Model 1 | Ref | 1.05 (0.83, 1.32) | 1.29 (1.03, 1.63) | 1.38 (1.08, 1.76) | 1.54 (1.17, 2.03) | 1.30 (1.15, 1.46) | <.0001 |
| Model 2 | Ref | 1.03 (0.82, 1.30) | 1.23 (0.98, 1.55) | 1.26 (0.99, 1.62) | 1.41 (1.07, 1.86) | 1.25 (1.11, 1.41) | 0.0003 |
| Model 3 | Ref | 1.02 (0.81, 1.29) | 1.21 (0.96, 1.52) | 1.23 (0.96, 1.57) | 1.30 (0.98, 1.73) | 1.20 (1.06, 1.35) | 0.004 |
| Stroke |  |  |  |  |  |  |  |
| Cases | 101 | 235 | 187 | 152 | 81 | 756 |  |
| Person-years | 69353 | 140907 | 119671 | 90035 | 50394 | 470359 |  |
| Model 1 | Ref | 1.22 (0.97, 1.55) | 1.24 (0.98, 1.58) | 1.46 (1.14, 1.88) | 1.47 (1.10, 1.98) | 1.19 (1.04, 1.35) | 0.009 |
| Model 2 | Ref | 1.21 (0.96, 1.52) | 1.21 (0.95, 1.54) | 1.39 (1.08, 1.79) | 1.40 (1.05, 1.89) | 1.16 (1.02, 1.33) | 0.02 |
| Model 3 | Ref | 1.21 (0.96, 1.53) | 1.21 (0.95, 1.55) | 1.40 (1.09, 1.81) | 1.41 (1.04, 1.90) | 1.16 (1.01, 1.33) | 0.03 |

Model 1: adjusted for age, sex, race, Townsend deprivation index, work schedules, and family history of CVD
Model 2: Model 1 + adjusted for BMI, smoking status, alcohol consumption, diet quality, physical activity, and history of hypertension, dyslipidemia, diabetes and depression
Model 3: Model 2 + adjusted for accelerometer-measured average sleep duration and sleep efficiency, self-reported insomnia symptoms, chronotype and daytime sleepiness, and clinically diagnosed sleep apnea based on ICD-10

**Supplemental Table 2.**
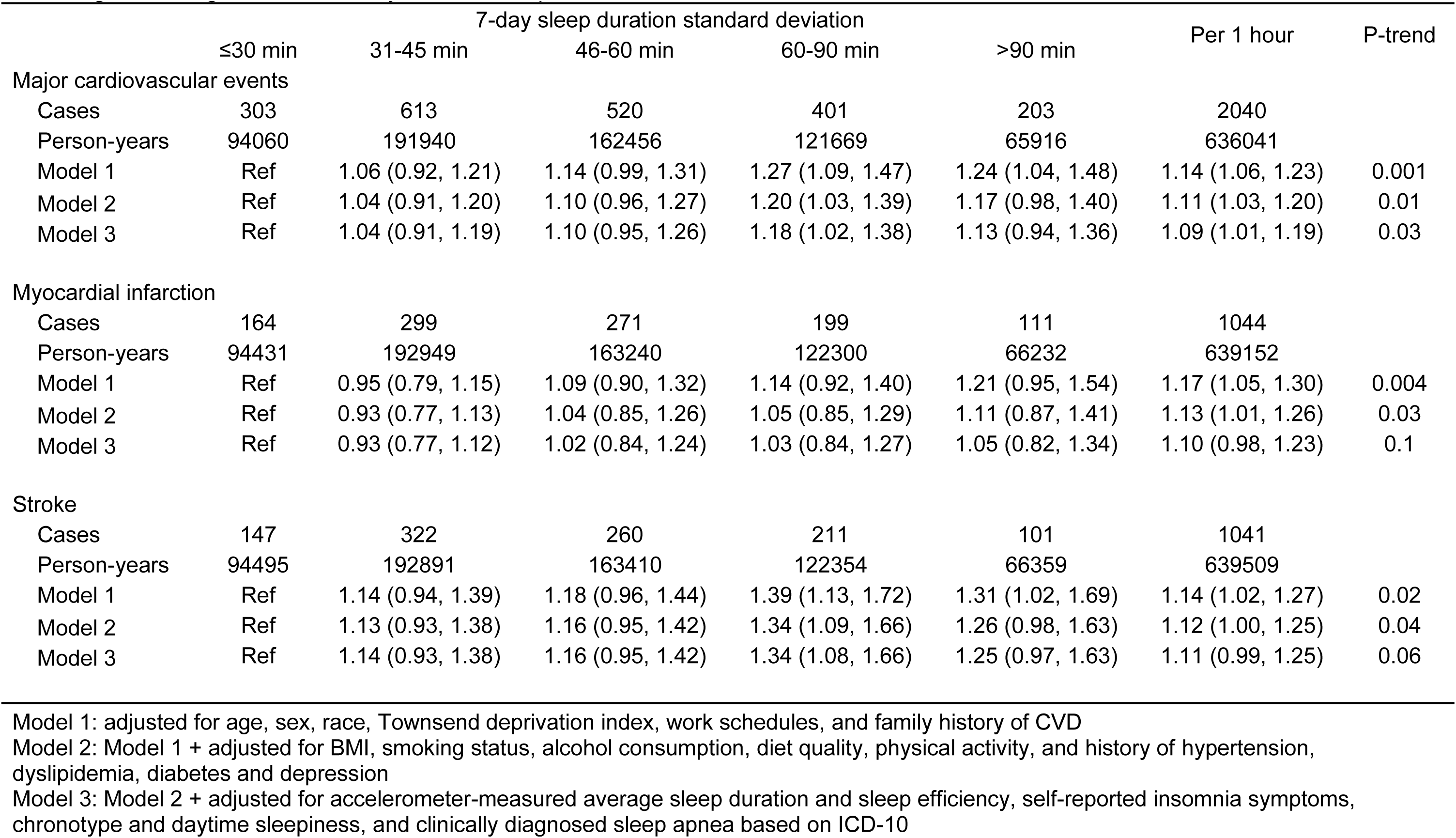
Associations of accelerometer-measured sleep duration irregularity with incident cardiovascular events in the UK Biobank, excluding cases diagnosed in the first year of follow-up.

**Supplemental Table 3.** Associations of accelerometer-measured sleep duration irregularity on weekdays with incident cardiovascular events in the UK Biobank.

|  | 7-day sleep duration standard deviation |  |  |  |  | Per 1 hour | P-trend |
| --- | --- | --- | --- | --- | --- | --- | --- |
|  | ≤30 min | 31-45 min | 46-60 min | 60-90 min | >90 min |  |  |
| Major cardiovascular events |  |  |  |  |  |  |  |
| Cases | 277 | 408 | 380 | 508 | 411 | 1984 |  |
| Person-years | 84861 | 122075 | 111436 | 138939 | 96557 | 553867 |  |
| Model 1 | Ref | 1.00 (0.86, 1.17) | 1.03 (0.88, 1.20) | 1.09 (0.94, 1.26) | 1.24 (1.07, 1.45) | 1.18 (1.09, 1.28) | <.0001 |
| Model 2 | Ref | 1.00 (0.85, 1.16) | 1.01 (0.86, 1.17) | 1.04 (0.90, 1.20) | 1.15 (0.99, 1.35) | 1.13 (1.04, 1.22) | 0.003 |
| Model 3 | Ref | 1.00 (0.85, 1.16) | 1.00 (0.86, 1.17) | 1.03 (0.89, 1.20) | 1.14 (0.97, 1.33) | 1.12 (1.03, 1.21) | 0.005 |
| Myocardial infarction |  |  |  |  |  |  |  |
| Cases | 141 | 203 | 178 | 263 | 205 | 990 |  |
| Person-years | 85221 | 122592 | 111986 | 139666 | 97108 | 556573 |  |
| Model 1 | Ref | 0.99 (0.80, 1.23) | 0.96 (0.77, 1.20) | 1.13 (0.92, 1.39) | 1.26 (1.02, 1.57) | 1.19 (1.06, 1.32) | 0.002 |
| Model 2 | Ref | 0.98 (0.79, 1.21) | 0.93 (0.74, 1.16) | 1.06 (0.86, 1.30) | 1.14 (0.92, 1.42) | 1.11 (1.00, 1.24) | 0.05 |
| Model 3 | Ref | 0.98 (0.79, 1.22) | 0.92 (0.74, 1.15) | 1.05 (0.85, 1.29) | 1.12 (0.90, 1.39) | 1.10 (0.98, 1.23) | 0.09 |
| Stroke |  |  |  |  |  |  |  |
| Cases | 139 | 214 | 212 | 253 | 214 | 1032 |  |
| Person-years | 85272 | 122666 | 111941 | 139716 | 97129 | 556725 |  |
| Model 1 | Ref | 1.04 (0.84, 1.29) | 1.13 (0.91, 1.40) | 1.06 (0.86, 1.30) | 1.24 (1.00, 1.54) | 1.18 (1.06, 1.31) | 0.002 |
| Model 2 | Ref | 1.03 (0.83, 1.28) | 1.11 (0.90, 1.38) | 1.02 (0.83, 1.26) | 1.18 (0.95, 1.46) | 1.14 (1.02, 1.27) | 0.02 |
| Model 3 | Ref | 1.03 (0.83, 1.28) | 1.11 (0.90, 1.38) | 1.02 (0.83, 1.26) | 1.17 (0.94, 1.45) | 1.14 (1.02, 1.27) | 0.02 |

Model 1: adjusted for age, sex, race, Townsend deprivation index, work schedules, and family history of CVD
Model 2: Model 1 + adjusted for BMI, smoking status, alcohol consumption, diet quality, physical activity, and history of hypertension, dyslipidemia, diabetes and depression
Model 3: Model 2 + adjusted for accelerometer-measured average sleep duration and sleep efficiency, self-reported insomnia symptoms, chronotype and daytime sleepiness, and clinically diagnosed sleep apnea based on ICD-10

**Supplemental Table 4.** Associations of accelerometer-measured sleep duration irregularity with risk of ischemic and hemorrhagic stroke in the UK Biobank.

|  | 7-day sleep duration standard deviation |  |  |  |  | Per 1 hour | P-trend |
| --- | --- | --- | --- | --- | --- | --- | --- |
|  | <30 min | 30-44 min | 45-59 min | 60-89 min | ≥90 min |  |  |
| Ischemic stroke |  |  |  |  |  |  |  |
| Cases | 118 | 266 | 211 | 186 | 98 | 879 |  |
| Person-years | 94519 | 192940 | 163443 | 122385 | 66379 | 639665 |  |
| Model 1 | Ref | 1.18 (0.95, 1.47) | 1.20 (0.96, 1.50) | 1.54 (1.22, 1.94) | 1.59 (1.21, 2.08) | 1.24 (1.10, 1.39) | 0.0003 |
| Model 2 | Ref | 1.17 (0.94, 1.46) | 1.17 (0.93, 1.47) | 1.47 (1.17, 1.86) | 1.51 (1.16, 1.99) | 1.21 (1.08, 1.36) | 0.001 |
| Model 3 | Ref | 1.18 (0.95, 1.46) | 1.18 (0.94, 1.48) | 1.48 (1.17, 1.87) | 1.52 (1.15, 2.00) | 1.21 (1.08, 1.37) | 0.002 |
| Hemorrhagic stroke |  |  |  |  |  |  |  |
| Cases | 44 | 105 | 81 | 55 | 24 | 309 |  |
| Person-years | 94519 | 192940 | 163443 | 122385 | 66379 | 639665 |  |
| Model 1 | Ref | 1.23 (0.87, 1.75) | 1.20 (0.83, 1.74) | 1.18 (0.79, 1.76) | 1.02 (0.62, 1.68) | 0.97 (0.78, 1.21) | 0.79 |
| Model 2 | Ref | 1.22 (0.86, 1.74) | 1.19 (0.82, 1.72) | 1.15 (0.77, 1.72) | 1.00 (0.61, 1.65) | 0.96 (0.77, 1.20) | 0.72 |
| Model 3 | Ref | 1.21 (0.85, 1.73) | 1.17 (0.81, 1.70) | 1.13 (0.75, 1.68) | 0.96 (0.58, 1.60) | 0.94 (0.75, 1.18) | 0.61 |

Model 1: adjusted for age, sex, race, Townsend deprivation index, work schedules, and family history of CVD
Model 2: Model 1 + adjusted for BMI, smoking status, alcohol consumption, diet quality, physical activity, and history of hypertension, dyslipidemia, diabetes and depression
Model 3: Model 2 + adjusted for accelerometer-measured average sleep duration and sleep efficiency, self-reported insomnia symptoms, chronotype and daytime sleepiness, and clinically diagnosed sleep apnea based on ICD-10

**Supplementary Table 5.** Subgroup analysis of the association between accelerometer-measured sleep duration irregularity and risk of myocardial infarction and stroke in the UK Biobank.

|  | Myocardial infarction |  |  | Stroke |  |  |
| --- | --- | --- | --- | --- | --- | --- |
|  | Cases/N | HR (95% CI)<sup>1</sup> | P-int | Cases/N | HR (95% CI)<sup>1</sup> | P-int |
| Age |  |  | 0.04 |  |  | 0.27 |
| <60 yrs | 228/31903 | 1.02 (0.82, 1.27) |  | 171/31903 | 1.02 (0.79, 1.32) |  |
| ≥60 yrs | 955/54316 | 1.29 (1.16, 1.44) |  | 1004/54316 | 1.20 (1.07, 1.34) |  |
| Sex |  |  | 0.87 |  |  | 0.04 |
| Men | 792/36651 | 1.21 (1.07, 1.36) |  | 653/36651 | 1.27 (1.11, 1.44) |  |
| Women | 391/49568 | 1.26 (1.06, 1.49) |  | 522/49568 | 1.04 (0.88, 1.23) |  |
| Race |  |  | 0.81 |  |  | 0.20 |
| White | 1144/83577 | 1.23 (1.12, 1.36) |  | 1145/83577 | 1.18 (1.07, 1.31) |  |
| Non-white | 39/2642 | 1.14 (0.70, 1.86) |  | 30/2642 | 0.76 (0.38, 1.54) |  |
| Average sleep duration |  |  | 0.46 |  |  | 0.0003 |
| <7 hrs | 483/29488 | 1.12 (0.99, 1.27) |  | 416/29488 | 1.01 (0.87, 1.17) |  |
| 7-8 hrs | 486/39890 | 1.24 (1.03, 1.50) |  | 513/39890 | 1.26 (1.05, 1.51) |  |
| >8 hrs | 214/16841 | 1.34 (0.95, 1.87) |  | 246/16841 | 1.73 (1.32, 2.27) |  |
| Family history of CVD |  |  | 0.005 |  |  | 0.75 |
| No | 363/34288 | 1.00 (0.82, 1.22) |  | 405/34288 | 1.15 (0.97, 1.34) |  |
| Yes | 820/51931 | 1.33 (1.19, 1.49) |  | 770/51931 | 1.18 (1.04, 1.34) |  |
| PRS<sup>2</sup> |  |  | 0.22 |  |  | 0.79 |
| Low | 198/28033 | 1.41 (1.14, 1.76) |  | 318/28033 | 1.00 (0.80, 1.25) |  |
| Medium | 358/28034 | 1.29 (1.08, 1.54) |  | 373/28035 | 1.34 (1.13, 1.58) |  |
| High | 599/28034 | 1.13 (0.98, 1.31) |  | 456/28033 | 1.14 (0.96, 1.35) |  |

<sup>1</sup>Adjusted for age, sex, race, Townsend deprivation index, work schedules, and family history of CVD
<sup>2</sup>Among 84101 participants with genotyping information with additional adjustment of the first three genetic principal components

**Supplemental Figure 1.**
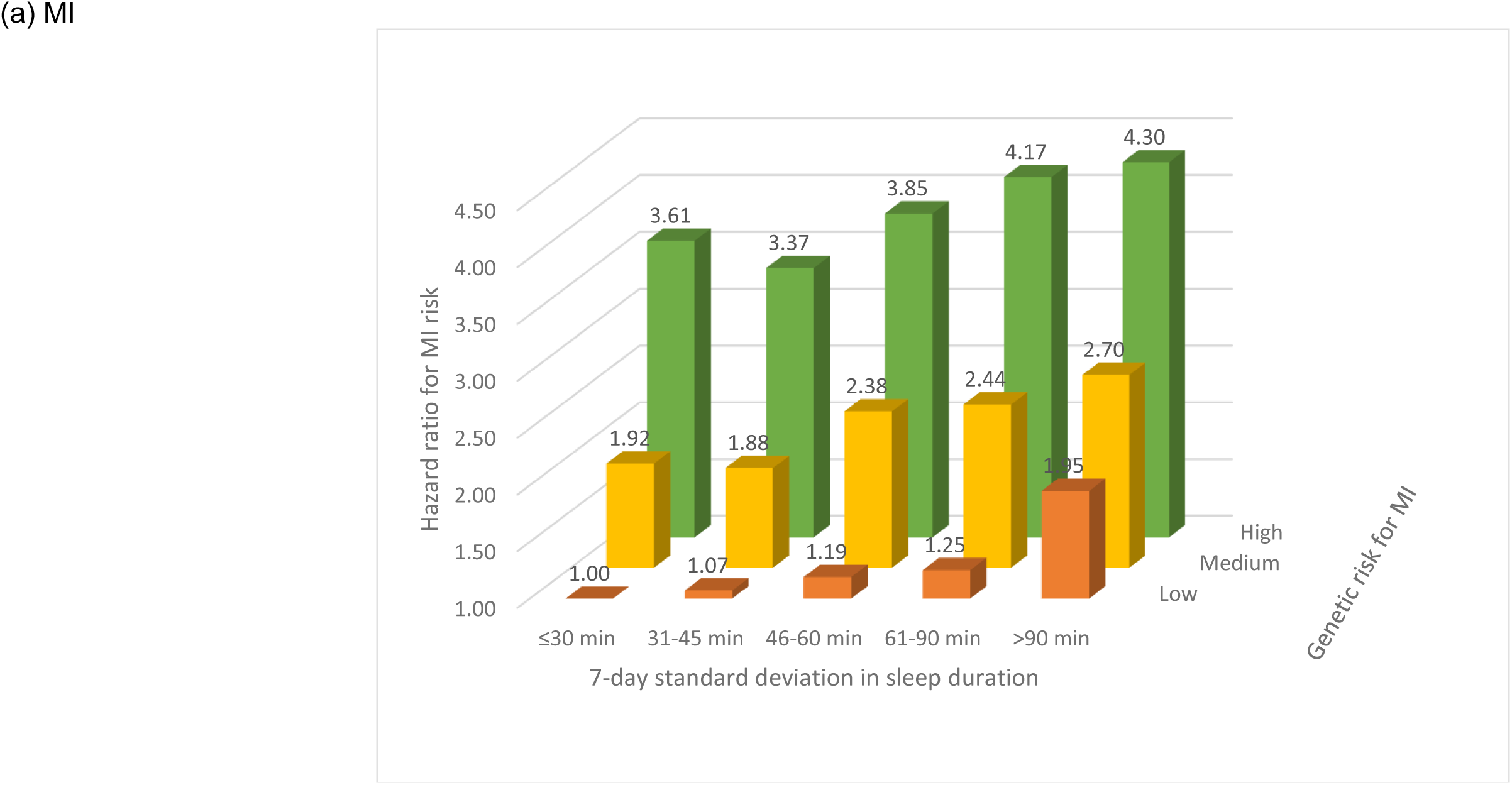
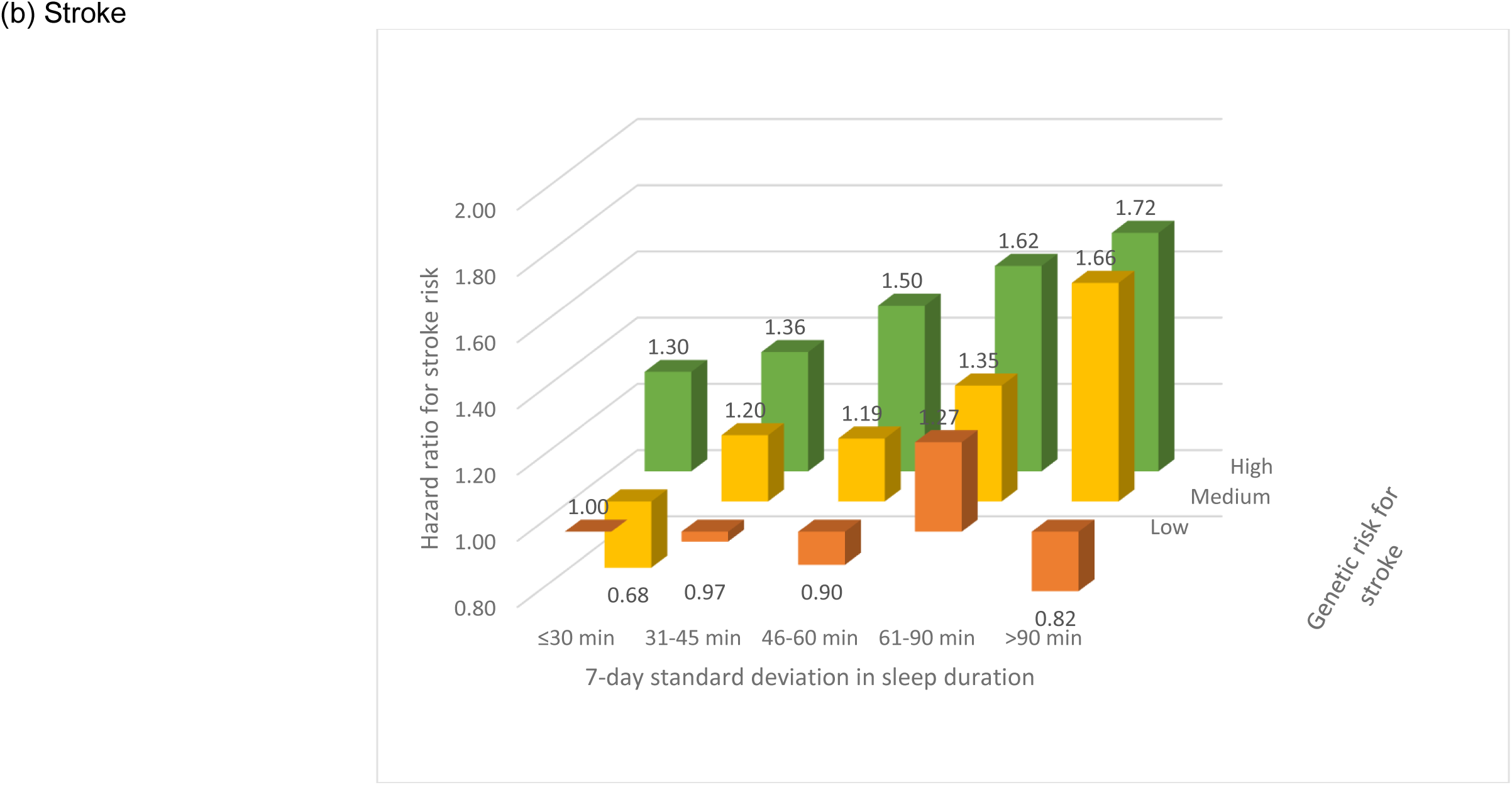
Risk of incident MI (a) or stroke (b) according to joint categories of sleep duration irregularity and polygenic risk score for MI or stroke. Estimates adjusted for age, sex, race, Townsend deprivation index, work schedules, family history of CVD, and the first three genetic principal components.

**Supplemental Figure 2.**
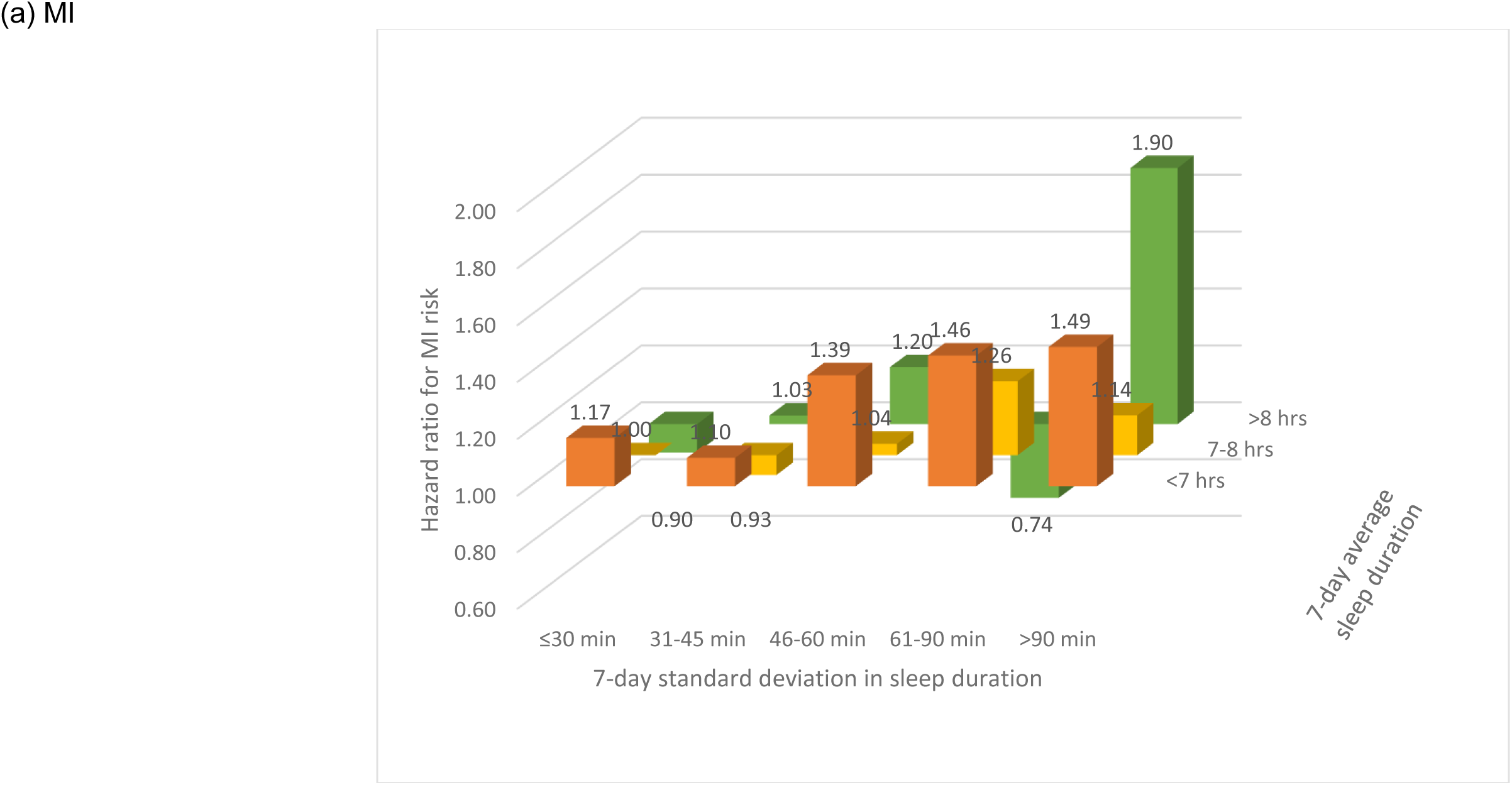
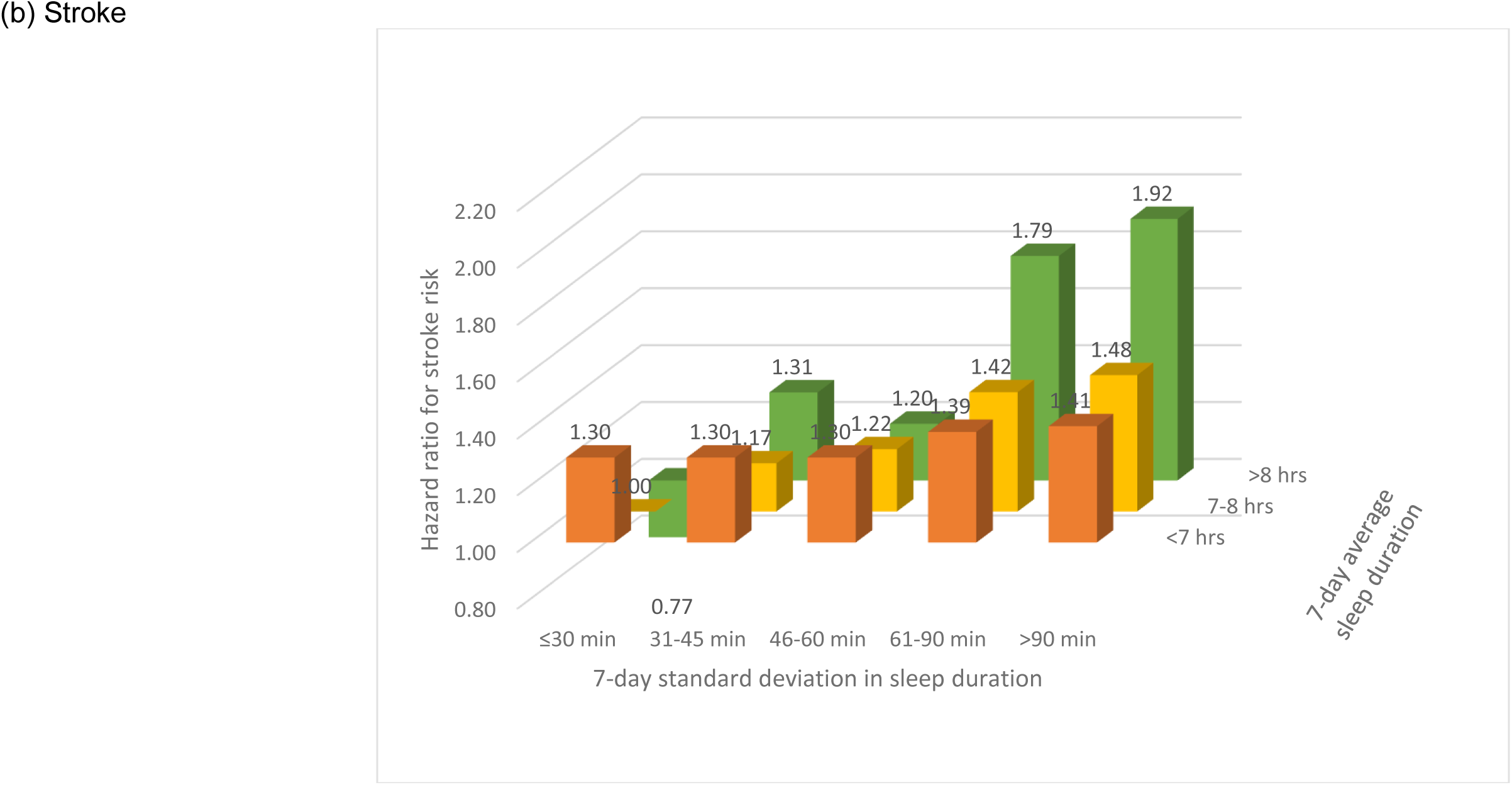
Risk of incident MI (a) or stroke (b) according to joint categories of sleep duration irregularity and average sleep duration. Estimates adjusted for age, sex, race, Townsend deprivation index, work schedules, and family history of CVD.

